# GENDER DIFFERENCES IN ANTIBIOTICS USE BEHAVIOUR AND ACCESS TO ANTIBIOTICS IN LOW- AND MIDDLE-INCOME COUNTRIES: A SCOPING REVIEW PROTOCOL

**DOI:** 10.1101/2023.06.06.23291021

**Authors:** Oluwafemi J. Adewusi, Rebecca Cassidy, Aaron O. Aboderin, Simon Bailey, Sarah Hotham

**Author notes:** **Author contributions** **Conceptualization**: Oluwafemi J. Adewusi **Methodology**: Oluwafemi J. Adewusi, Rebecca Cassidy, Sarah Hotham, **Supervision**: Rebecca Cassidy, Aaron O. Aboderin and Sarah Hotham **Writing (Original draft)**: Oluwafemi J. Adewusi **Writing (review and editing)**: Rebecca Cassidy, Simon Bailey, Aaron O. Aboderin and Sarah Hotham. **Approval of manuscript**: All the authors. **Funding** The study received no funding.

## Abstract

**Background:** In low-and middle-income countries (LMICs), the persistent lack of access and high inappropriate use of antibiotics, which are fuelled by gender-related factors continue to facilitate antimicrobial resistance which in turn reduces the capacity to treat infectious diseases. However, there is lack of sufficient clarity on the nature and extent of gender influence on access to antibiotics and antibiotic use behaviour. This proposed study aims to systematically review available literature in order to map out the nature and extent of evidence on gender differences and related factors influencing antibiotics use behaviour and access to antibiotics in LMICs.

**Methods:** This scoping review will be conducted using the Preferred Reporting Items for Systematic Reviews and Meta-Analyses (PRISMA) guidelines for scoping reviews. The MEDLINE, PsycINFO, African Journal Online (AJOL), Web of Science and CINAHL databases will be searched for peer-reviewed articles and relevant electronic grey literature will also be included in the study. A predefined excel spreadsheet will be utilized for data extraction and analysis. Findings will be presented in narrative summary and tables.

**Conclusion:** It is expected that this study will identify knowledge and gaps on gender contributory factors to antibiotics use and access to antibiotics. These will contribute to understanding gender health inequalities and areas for further research on gender mainstreaming in antimicrobial stewardship efforts in the LMICs. This study findings will be disseminated through presentations in scientific conferences and publications in peer-reviewed journals.

**Scoping review registration with Open Science Framework:** Registration DOI: https://doi.org/10.17605/OSF.IO/N5W8E

## INTRODUCTION

The rates of antimicrobial resistance (AMR) are increasing worldwide especially in low-and middle-income countries (LMICs) (1), resulting into the use of broad-spectrum antibiotics including those considered as last resort for the treatment of bacterial infections (2). This leaves clinicians with limited therapeutic options for the treatment of infections, and also potentially signifies future inability to treat infections. Apart from the clinical implications, AMR will significantly increase healthcare cost, poverty (3) and the World Bank predicted a 100 trillion USD global production loss by 2050 if there are no AMR policy interventions (4).

Notably, AMR is fuelled largely by modifiable human behaviour, primary of which is inappropriate or irrational antibiotics use (1). As evidenced from growing literature, antibiotics use behaviour change is complex and non-linear (1). This is reflected in the dynamics around self-medication (5), non-adherence to medical prescription (6) and use of antibiotics for viral infections (7). The complexities in antibiotic use behaviour are fuelled by factors such as the lived experience of illness, social perception of antibiotics, access and cost of antibiotics, and societal expectation of disease treatment (8). Previous studies also showed that inappropriate antibiotics behaviour in the LMICs is associated with income, level of education, perception of illness and antibiotics, sex and other cultural related issues including gender (6)(5)(9).

Despite the increasing use of antibiotics, there are reported inequity gaps in access to this medicine. The Centre for Disease Dynamics, Economics and Policy (CDDEP) reported in 2019 that LMICs had limited access to antibiotics compared to their HICs counterparts and these were attributed to patients’ financial incapability, limited resource allocations to health by the government and poor local and international drug logistics and infrastructure. The report revealed that dying from treatable microbial infections due to lack of access to adequate antibiotics indirectly fuel antimicrobial resistance through death from treatable bacterial infections which further create a cycle of ineffective antibiotics use (10).

The contributions of gender to health are well documented, playing crucial direct and indirect dynamic roles in disease perceptions, prevention practices, health-seeking behaviour and treatment, and access to healthcare (11)(12). Although the pattern of gender inequalities varies according to context, women are largely affected due to the patriarchy systems in most LMICs. They often lack economic resources, incapable of independently taking health decisions and hence, unable to access needed healthcare (13). For instance, with 26% poverty incidence and 17% access to healthcare services among women-headed households compared to 24% poverty incidence and 41% access to healthcare services among male-headed households in Nigeria (14)(15), the risk of exposure to resistant infections and inappropriate antibiotics use behaviour are higher for women and needless to say that this further sustains the vicious cycle of poverty and inequalities (16).

Results from past studies also indicated that women were 1.65 times more likely to be infected with resistant pneumoniae infection largely because of motherhood roles (17) and that poor hand hygiene practices during childbirth also predispose them to resistant infections (11). In addition, experience of being labelled promiscuous and ‘wayward’ by the health workers when suffering from sexual health and urinary tract infections could make women resort to self-medication with antibiotics (18).

In spite of the reportedly high prevalence of inappropriate use of antibiotic across different categories of the population in the LMICs, most of these countries lack policies or guidelines strategically addressing access to antibiotics and inappropriate use behaviour with consideration to gender. Increasing access to antibiotics will not only reduce death due to bacterial infections, it is will also prevent AMR. Hence, the importance of addressing gender inequalities in access to this medicine as well as in antibiotics use behaviour.

Most existing reviews on human antibiotics behaviours focused on physicians’ prescribing behaviour (19)(20) and other studies examined users’ behaviour (21)(9) with no gender reference thus creating a gap in the knowledge of the gender-related determinants of antibiotics use. Similarly, there is little evidence on how gender factors influence access to antibiotics, and it is against this backdrop that we aimed to conduct a scoping review. We intend to utilize this scoping review to examine the range and characteristics of evidence available in the literature as well as knowledge gap on the gender differences in antibiotics use behaviour and access to antibiotics in the LMICs.

### Main objective

The overarching aim of this review is to systematically identify the nature, themes and knowledge gaps on antibiotics use behaviour and access to antibiotics with reference to gender in LMICs.

### Review questions

1. What gender differences in antibiotic use exist in the LMICs?
2. What factors contribute to the gender differences in antibiotic use behaviour in the LMICs?
3. What gender differences in access to antibiotic exist in the LMICs?
4. What factors contribute to the gender differences in access to antibiotic in the LMICs?

**Table 1:**
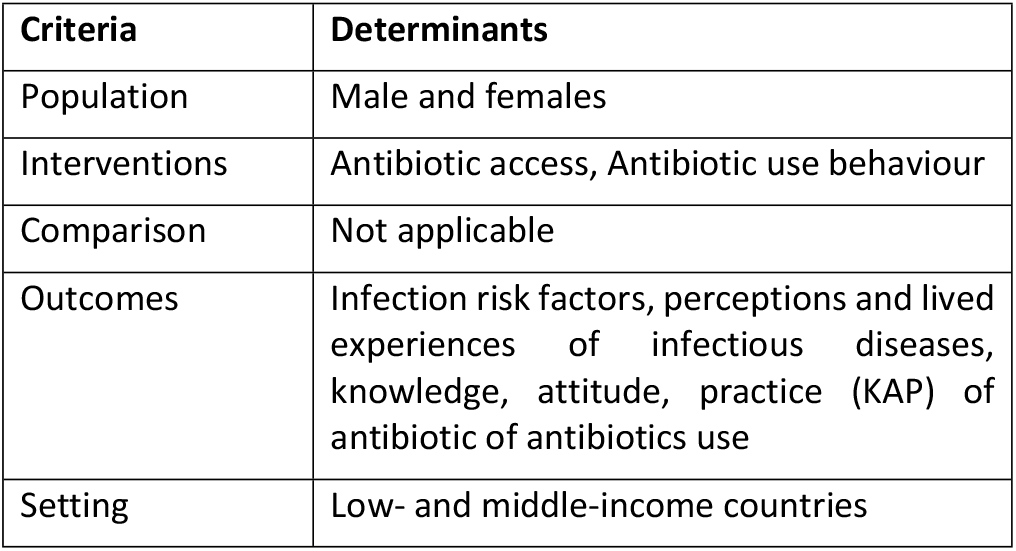
Guiding PICOS framework for the study question.

## METHODOLOGY

### Protocol and registration

The study protocol was registered with the Open Science Framework with registration digital object identifier: https://doi.org/10.17605/OSF.IO/N5W8E.

### Search strategy

This review will be conducted to follow the Preferred Reporting Items for Systematic Reviews and Meta-Analyses (PRISMA) guidelines for scoping review (22). The review will be conducted between June and December 2023. The MEDLINE, PsycINFO, African Journal Online (AJOL), Web of Science and CINAHL databases would be searched for peer-reviewed primary studies relevant to the study research questions. Grey literature such as technical reports from organizations working on antibiotics use in the LMICs will also be included. Additional publications will be checked from the reference lists of relevant articles using snowballing technique. Authors of relevant papers would also be emailed for access to non-readily available and additional relevant papers. No date or periodic coverage limitation would be applied to the search.

### Search terms

In this study, ‘access’ refers to affordability and availability of antibiotics. These are hinged on two of the four blocks within the WHO framework for access to medicine. These two blocks (affordable prices and reliable health and supply systems) were selected because they relate more with the consumers as other blocks (Rational selection and sustainable financing) rely largely on the health system (23). Therefore, this study considers financial and economic capability to purchase as well as availability of needed antibiotics.

Keywords to be used for the search will include ‘antibiotic use’, ‘antimicrobial use’, ‘respiratory tract infections’, ‘diarrhoea’, ‘anti-bacterial/anti-infective agents’, ‘prudent antibiotic use’, ‘judicious antibiotic use’, ‘self-medication’, ‘practice’, ‘access*’, ‘afford*, ‘gender’, ‘male’, ‘female’, ‘Africa’, ‘sub-Sahara’, ‘Asia’, ‘low-and middle-income countries’.

### Inclusion and exclusion criteria

The literature search will be limited to literature with the following criteria:

- primary peer and non-peer reviewed publications in English language
- studies reporting antibiotics use among males and females
- studies reporting antibiotics access among males and females
- studies conducted in LMICs (as defined by the World Bank (24)
- all study designs

Studies with the following criteria will be excluded:

- Studies published as conference abstracts reviews, editorials or letters to the editor on AMU
- studies reporting antibiotics use in animals
- studies describing antibiotics prescribing behaviour and factors
- studies generally reporting antibiotics use without reference to males or females
- Studies conducted in high income countries

**Figure 1:**
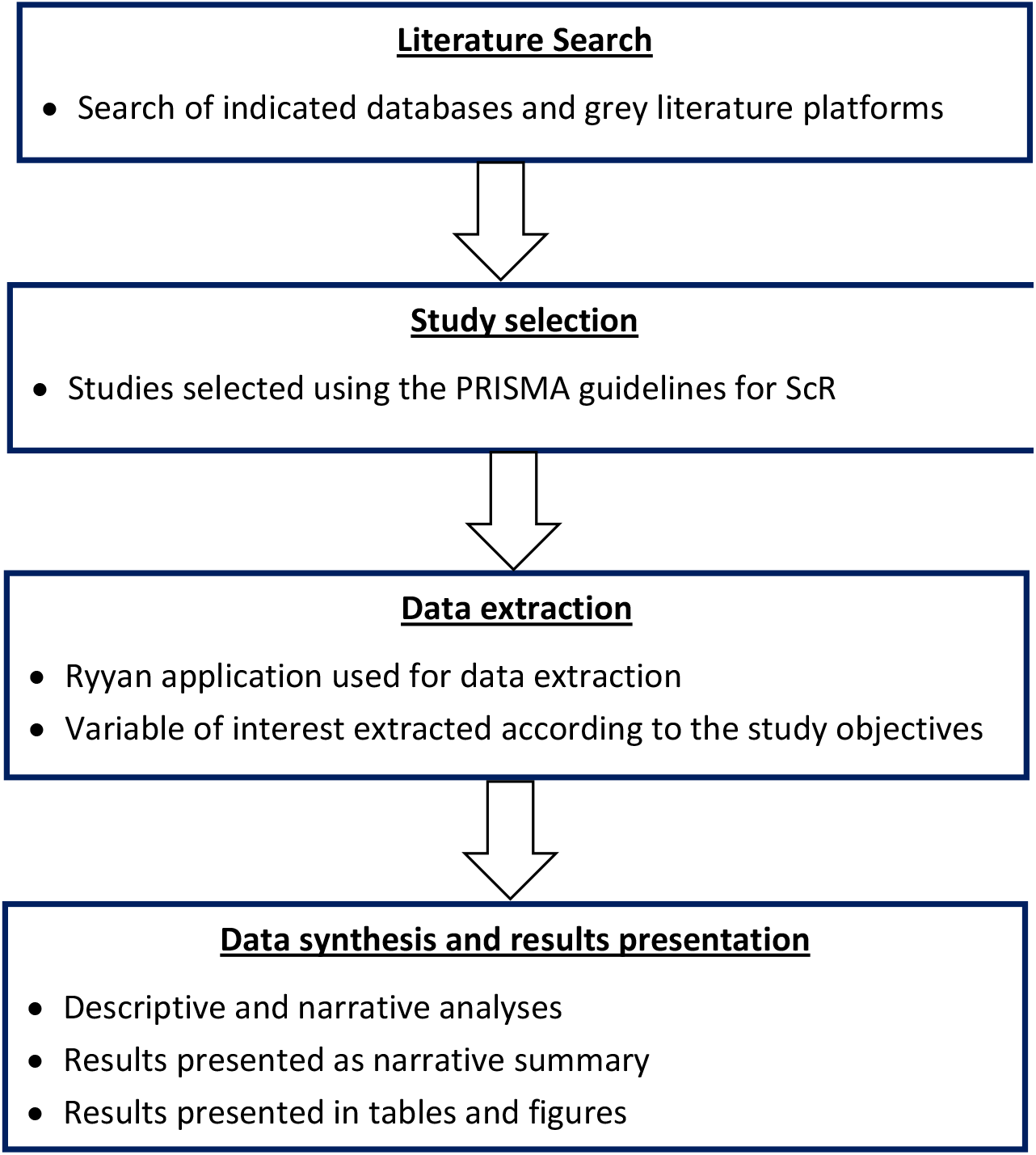
Flow diagram for the scoping review methodology on gender differences in antibiotics use behaviour and access to antibiotics in LMICs.

### Data extraction/variables of interest

Data extraction will be carried out from all eligible articles using a predefined excel sheet, by the first author with supervision from all the other senior authors. The following variables will be extracted from the reviewed studies; title, author, year of publication, target population and characteristics, country of study, study setting, study design, sample size, gender factors affecting antibiotic access, gender factors affecting antibiotic use practices and behaviour.

### Data charting and results presentation

The extracted data will be organized in an excel spreadsheet and descriptive analysis will be carried out to characterise the studies. A thematic analysis would also be carried out on the review findings, and these will be presented in tables and charts where appropriate. The study findings will be summarized in narrative that synthesize the results across the data variables, themes and knowledge gaps with possible implications for further studies.

### Ethical consideration

This would not be applicable to this study.

## CONCLUSION

While there are increasing studies outlining the factors influencing access and antibiotics use behaviour in LMICs, there are limited evidence the gender dynamics of these factors. This scoping review will map out evidence on the gender differences on access and antibiotics use behaviour in LMICs. The dissemination plan for the findings of this review include seminar presentations at the University and local public health institutions, conference presentations and publication in peer-reviewed journals.

## Data Availability

Research data will be submitted as supplementary materials to be published when the study is completed.

